# Online psychoeducation and assessment for borderline personality disorder as a first step of care: A pilot study assessing safety, feasibility, and mechanisms of change

**DOI:** 10.64898/2026.05.29.26354218

**Authors:** Lois W. Choi-Kain, Destiny J. Crisp, Sam Mermin, Grace E. Murray, Julia B. Jurist, Sara R. Masland, Margaret Mosby, Laura T. Germine, Boyu Ren

## Abstract

**Background:** Treatment guidelines for borderline personality disorder (BPD) recommend assessment, diagnosis, and psychoeducation. We report on the feasibility and safety of a randomized controlled trial protocol of online psychoeducation, assessment, and personalized feedback as an immediate first step of care for BPD.

**Methods:** Newly diagnosed participants were randomized to receive 10 videos about BPD or general mental health for two weeks. Half the participants receiving BPD videos were randomized to receive personalized feedback on changes in symptom ratings and cognitive performance. Ecological momentary assessment (EMA) evaluated interpersonal interactions, emotions, and behaviors for 30 days. BPD symptoms, depression, and personality functioning were assessed at baseline, after videos, after feedback, and one month later.

**Results:** Eighty-two participants were randomized into three conditions that did not differ significantly in terms of demographics or baseline variables. Dropout occurred for 32.9% of the sample. No differences in rate of emergency room visits, hospitalizations, or other escalations in level of care were reported among groups. Satisfaction was higher for those receiving psychoeducational videos about BPD. Improvement in BPD knowledge in the psychoeducation conditions was significantly greater than the control condition. No statistically significant differences were found regarding reduction of BPD symptoms. The psychoeducation with feedback arm showed significantly greater improvements in self-impairment compared to controls with medium effect size at the final timepoint. Modeling of the relationship between time spent alone and BPD symptoms showed a positive correlation in the control condition, but in the group receiving both psychoeducation about BPD and feedback, this relationship was negative.

**Conclusion:** Online psychoeducational videos and assessment were safe, feasible, and acceptable to participants with newly diagnosed BPD. Psychoeducation with personalized feedback appears to be more effective than either BPD or general psychoeducation alone in improving deficits in self-functioning, which may relate to an increased capacity to be alone with fewer symptoms.

The protocol was registered with ClinicalTrials.gov (NCT05358925, https://clinicaltrials.gov/study/NCT05358925) on April 28^th^, 2022.

## Introduction

Borderline personality disorder (BPD) is a prevalent and severe mental illness characterized by frantic efforts to avoid abandonment, unstable sense of self, emotional dysregulation, and self-destructive impulsivity [1]. Associated with significant psychiatric and medical morbidity, as well as social adversity, its personal and societal costs are high [2–3]. These costs result not only from direct care, but also from damaging reckless impulsivity, lost productivity, and informal or community care compounding to 62,260 euros (i.e., approximately $72,000) per patient reported annually [4]. Fortunately, people with BPD respond to a variety of treatments tailored to address its core features [5–6]. Yet such treatment is rarely the first step of care [7]. Even in countries best resourced with specialized psychotherapies for BPD, only a minority of patients will receive psychotherapy immediately after diagnosis due to the restricted supply of trained professionals available to provide them [8]. Worldwide, the availability of recommended therapies for BPD is grossly insufficient, leaving people with a disorder defined by frantic efforts to avoid being abandoned at risk for self-injury and destructive interpersonal dynamics without alternatives for care. Our own study using publicly available data for healthcare workforce combined with records of specialist therapy certification bodies in 22 countries indicated that no country possessed an adequate supply of certified specialists to meet demands [8]. Even in nations with enriched specialist personality disorder services such as the Netherlands, psychotherapy is rarely the first step of care, despite being the primary recommended treatment for the diagnosis of BPD [7]. Most clinicians diagnosing BPD are often left without pragmatic tools to immediately initiate tailored treatment.

Scalable, less specialized and intensive steps of care are needed to fill the gap between the urgent needs of patients diagnosed with BPD and the supply of guideline-informed care. Recently updated American Psychiatric Association (APA) Guidelines [5] recommend routine diagnostic assessment, disclosure, and psychoeducation, followed by psychosocial interventions targeting core mechanisms of the disorder. The rigorous systematic review that informed the revision of APA guidelines from its original 2001 version [9] found that numerous treatments tested for BPD, including treatment as usual (TAU), were effective in reducing its symptoms. This review reported low-certainty evidence that specific therapies, such as dialectical behavioral therapy (DBT [10]), mentalization-based treatment (MBT [11]), transference focused psychotherapy (TFP [12]), or schema therapy (ST [13]), were superior to TAU [14]. In a prior meta-analytic review, Cristea and colleagues reported small between group effects for DBT and psychodynamic therapies compared to TAU, despite the relatively more intensive format of these approaches, stating that length and intensity of treatment did not significantly influence outcome [6]. Consequently, the APA guidelines revision committee focused its recommendations on components of general psychiatric management that most practitioners can integrate into their treatment of people with BPD, such as routine diagnostic assessment and psychoeducation, in addition to psychosocial treatment targeting key mechanisms such as emotion dysregulation and interpersonal instability [5].

Studies of briefer variants of DBT [15] and MBT [16] demonstrate non-inferiority of variants that are half the length as the original formats, supporting the notion that duration of intervention is not necessarily a significant predictor of better outcomes [6]. However, abbreviated DBT skills training is often offered as an intervention for repeated self-injury without adequate evidence of effective dosage. A large pragmatic randomized trial of over 18,000 patients identified with recurrent self-injury receiving treatment in real world conditions within a large insurance-based healthcare program compared a low dose of online DBT skills training to TAU and a collaborative care approach [17]. This low dose of online DBT skills training was associated with a 30% increased risk for self-injury relapse and increased utilization of emergency room visits compared to TAU and a collaborative care approach [17]. Low dose implementation of intensive manualized psychotherapies appears common in clinical practice, but effectiveness in terms of retention in treatment and self-destructive problems appears influenced significantly by adherence in DBT, as an example [18].

Psychoeducation, as a preliminary step of less intensive treatment, is a practical starting point of care immediately after diagnosis of BPD. Brief, less resource-intensive interventions that provide psychoeducation delivered by research staff [19], in group therapies [20], and online [21] are found safe, feasible, and effective in reducing BPD symptoms more than waitlists. A six-session psychoeducational group informed by Good Psychiatric Management (GPM [22]) integrated information about BPD’s symptoms, interpersonal hypersensitivity, etiology, course, co-occurring disorders, and treatments resulted in greater reduction in total BPD symptoms measured by the ZAN-BPD scale than a waitlist comparison [20]. GPM’s interpersonal hypersensitivity model identifies intolerance of aloneness as an underlying mechanism driving dependency and reactivity, explaining that clinical presentation shifts when feeling connected with caring others, which can stabilize symptoms, whereas feeling threatened in terms of rejection or abandonment can trigger self-destructive, impulsive symptoms and suicidality [23]. Forty-six percent of participants receiving this GPM psychoeducational group achieved over 50% reduction of BPD symptoms from baseline, compared to 3% of the waitlist group. Intervention effects remained stable for two months after the group ended, suggesting that the impact of the group cannot solely be attributed to ameliorating loneliness through social connections available during group therapy. The effectiveness of this brief psychoeducational intervention, which oriented patients to their problems of interpersonal hypersensitivity and basic knowledge about their diagnosis, reflects the potential to relieve patients of significant proportion of their BPD when well-known empirically supported options are not immediately available [20, 24].

Digital or online applications could vastly amplify the scalability of psychoeducational interventions while providing increased flexibility, time, and space for patients to review and learn from materials at their own pace. Eight randomized control trials (RCTs) have evaluated digital interventions to date, many of which integrate psychoeducation. Of these, three RCTs employed an active comparison condition, as opposed to a waitlist, reporting the smallest differences between groups [25–28]. The largest of these RCTs (*N* = 580) examined *Priovi*, a digital ST intervention [28]. Offered twice a week with daily prompts for three to four months, *Priovi* combines psychoeducation about BPD with lessons drawn from schema focused therapy. *Priovi* combined with treatment as usual (TAU) yielded greater reduction in BPD symptoms, suicide attempts, depression, and anxiety than TAU alone. The between groups effect size in reducing BPD symptoms was small (*d* = 0.24) after three months of treatment. In addition, the generalizability of these findings requires further study. *Priovi* was tested in Germany where impressive gross domestic product allocations to national healthcare make psychiatric treatment available to most citizens, rendering a TAU of high quality when compared to other locales [29].

Online video prescriptions, which educate patients about their diagnosed conditions and available treatments, are routine across healthcare [30–31]. A Pew Research study reported that 72% of adults use the internet to learn about their health conditions [32]. In a qualitative study of the use of YouTube videos for psychoeducation, people with lived experience reported that online resources about BPD catalyzed recovery by increasing self-awareness, reducing self-stigma, and improving clarity in one’s sense of self [33]. Only one RCT to our knowledge has compared the effects of administering psychoeducational videos (on compact disc players) about DBT and MBT skills combined with live mentoring/coaching and care as usual, which reduced anxiety and self-harm more than care as usual alone [34].

Online video psychoeducation provides flexible, scalable means of reaching more patients without dependence on clinician face-to-face time or training. Providing tools for clinicians to conduct online assessments to guide care could enable personalization for individual patients in combination with more general diagnostically oriented psychoeducation. Studies of feedback of symptoms and clinical response to treatment show small effects on treatment outcomes in mental healthcare [35–36]. Structured measurement of symptoms can be helpful to both clinicians and patients, yet a minority of clinicians in the community utilize standard measurement tools [37].

In this report, we describe the results of a pilot RCT to 1) assess the safety and feasibility of prescribing 10 online educational videos about BPD as a scalable intervention to reduce BPD symptom severity after diagnostic disclosure as a first step of care; 2) evaluate the feasibility of an online clinical measurement protocol that incorporates standard self-assessment, EMA, and neuropsychological testing in participants with BPD to assess symptom severity and change, improvements in BPD knowledge, cognitive functioning, and underlying mechanisms of change; and 3) assess the added effects of an automated descriptive feedback report as a means of personalizing the intervention to the individual characteristics of the patient, which we hypothesize will increase self-clarity and awareness of fluctuations in BPD symptoms, depression, anxiety, and cognitive functioning. We hypothesized that psychoeducational videos that provide a basic orientation to BPD (i.e., its symptoms, course, and treatments) with online assessment and feedback would be safe, feasible, and acceptable. While this pilot study was not powered to assess efficacy, we assessed the relative effect that psychoeducation about BPD had on BPD symptoms, general personality functioning, and depressive symptoms compared to those in the control condition who received more generic psychoeducation on mental health. We also evaluated whether this effect is associated with increased knowledge about BPD. Lastly, we tested the feasibility of using EMA to track changes in social interactions and time spent alone over the course of the intervention period to assess if the intervention affected symptoms in the context of daily fluctuating social functioning, which may indicate a mechanism of change.

## Materials and methods

### Participants

Participants were adults in the United States who had been diagnosed with BPD within six months prior to study enrollment and had never received evidence-based, BPD-specific treatment, such as DBT, MBT, or TFP. Recruitment sites included five inpatient units at McLean Hospital, a central participant recruitment website for the larger Mass General Brigham (MGB) system within which McLean Hospital operates, and online advertisement via ResearchMatch, ClinicalTrials.gov, Prolific, and Instagram. Potential participants from inpatient units were identified via results of routine screening on the McLean Screening Instrument for Borderline Personality Disorder (MSI-BPD [38]) and consultation with clinical staff regarding their appropriateness and consent to be approached for research. Eligibility was then determined through a brief, in-person screening with a research assistant. Participants who expressed interest upon viewing online advertisements completed an online screening survey and were invited to complete a full screening with a research assistant on Zoom if their survey results indicated potential eligibility. If eligible, participants completed written informed consent forms. Participants were considered eligible if they reported either a diagnosis from a mental health professional or a self-diagnosis. Inclusion criteria included: (a) reliable access to a smartphone with a data plan for the duration of the study, (b) ability to speak and understand English, (c) age 18 or older, (d) diagnosis of BPD within the past six months, and (e) awake and able to complete daily EMA surveys between 9am and 9pm Eastern Standard Time. Initially, recruitment was restricted to those within Massachusetts but later expanded nationally to broaden enrollment. Additionally, recruitment originally targeted individuals diagnosed with BPD within the past three months but later included diagnoses within the past six months. We later also allowed participants with prior exposure to, but not a full course, of DBT, MBT, or TFP to extend eligibility. This enabled the inclusion of those who had received DBT or MBT group therapy, but no participant completed a full course of adherent therapies of these types before or during their participation in this study. Individuals with significant cognitive disability were excluded when impairment interfered with demands of the study protocol. Other exclusion criteria included acute mania, acute psychosis, eating disorder symptoms threatening medical stability, and acute problematic substance use. The recruitment flow is represented in the CONSORT diagram (Fig 1).

**Fig 1.**
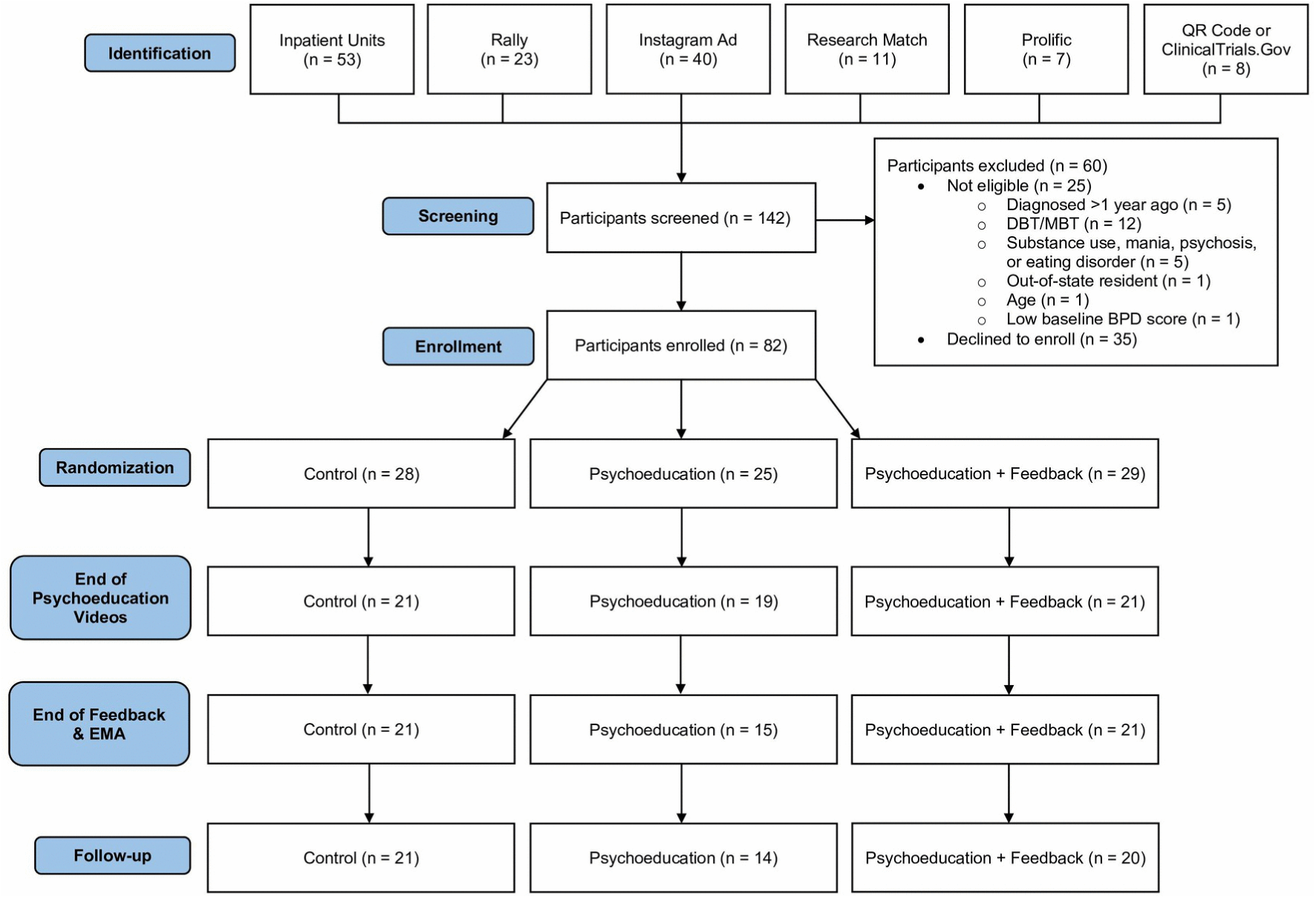
CONSORT flow diagram. Exclusion criteria were later expanded to include out-of-state participants and also include partial exposure to DBT, MBT, or TFP to extend eligibility. Twelve participants were excluded before this change was implemented.

The study was approved by the MGB IRB (2022000892). The protocol was registered with ClinicalTrials.gov (NCT05358925). Recruitment and data collection occurred between June 2022 and November 2024. The protocol is published in a separate report [39].

### Design

Our block randomization assigned participants to three groups: one received BPD psychoeducational videos and feedback (“psychoeducation + feedback group”); one received BPD psychoeducational videos without feedback (“psychoeducation group”); and one received generic mental health videos and no feedback (“control group”), which deviated from the original Sequential Multiple Assignment Randomized Trials design, which was no longer feasible given our smaller-than-expected sample size. This decision was made to maximize the likelihood of matched groups, instead of employing a second randomization based on treatment response. The computer-generated block randomization was weighted to assign 2/3 of participants to the psychoeducation condition at the first randomization point and to evenly divide those participants between feedback and no feedback at the second randomization point, creating three groups of equal size. Randomization factored in participants’ gender and baseline BPD symptom severity (divided into “high” and “low” based on a pre-determined threshold of 2.67 on the BSL-23) to balance these variables across conditions. Participants and the statistician were blinded in the randomization assignment. Limited blinding to treatment condition assigned could be achieved because participants could determine if their videos were about BPD or general mental health and whether they received feedback.

### Interventions

#### Psychoeducation

After enrolling and completing the baseline survey, all participants were sent links to short educational videos on ten consecutive weekdays via smartphone text messaging. Participants in the two psychoeducation groups (psychoeducation and psychoeducation + feedback) received a series of videos designed for this study as a cohesive introduction to BPD for newly diagnosed patients. Collaborating with the production team of the YouTube channel *BorderlinerNotes* lead by Rebbie Ratner, a lived experience expert, we produced 10 video prescriptions of each approximately six minutes in length. The contents of the videos were modeled from Ridolfi and colleagues six-session GPM psychoeducation group [20]. The first five videos delivered over the span of the first week reviewed basic facts of BPD, including its symptoms, prevalence, etiology, course, self-destructive behaviors and their management, and co-occurring disorders. Five more videos were administered the following week and educated participants about common factors in effective treatment for BPD, specific treatment approaches, the lack of FDA approved pharmacological treatments, and a summary with top 10 tips for living with BPD (S1 Table). Videos on specific treatment approaches presented various formulations of BPD in terms of emotion dysregulation and unstable mentalization. One video described the GPM [22] conceptualization of BPD as a disorder of interpersonal hypersensitivity. In total, the 10 videos were 66 minutes in length. Lived experience experts, both in recovery and carers, provided feedback on the videos. The control condition watched general mental health videos of similar length about topics other than BPD (e.g., depression, anxiety, sleep, self-care). Taken from a publicly available webinar series developed by McLean Hospital, the control videos were matched with the psychoeducational videos on both the institutional prestige and credibility of expert clinicians.

#### Feedback

All participants were asked to complete a series of self-report symptom measurements and cognitive tests at four time points over 60 days and a shorter battery of cognitive tests each day for the first 30 days (Fig 2). After the second time point, between day 16 and day 22, after viewing all the prescription videos, participants in the psychoeducation + feedback condition received an email summary of the changes in their symptoms and cognitive testing performance during the first period of the study (day 1 – day 15). These summaries were provided in a standardized PDF with graphical depictions of participants’ baseline and time point B scores on four symptomatic measures: overall borderline symptoms (with endorsement levels depicted for each individual item of the Borderline Symptom List – Brief Version (BSL-23 [40]); depressive symptoms (Patient Health Questionnaire (PHQ-9 [41]); loneliness (Loneliness Scale – Short Form (LS-3 [42]); and personality functioning (Level of Personality Functioning Scale (LPFS-BF [43]). After the symptomatic measures, a bar graph showed the total number of supportive connections and stressful interactions the participant had reported between baseline and time point B. Finally, the four cognitive tests depicted the results for sustained attention, response inhibition, processing speed, and short-term memory. The feedback was also accompanied by brief descriptions of each measure and the conceptual meaning of scores. This feedback format was designed to emphasize change in score rather than overall levels, aimed to help the participant gain self-awareness by recognizing changes in their own experience. At the end of the feedback, a “summary” section listed areas in which the participant had shown the greatest improvement, areas in which they had remained stable, and areas in which their symptoms had worsened, which was labelled “areas needing more support”. A sample of automated feedback is depicted in supplement S1 File.

**Fig 2.**
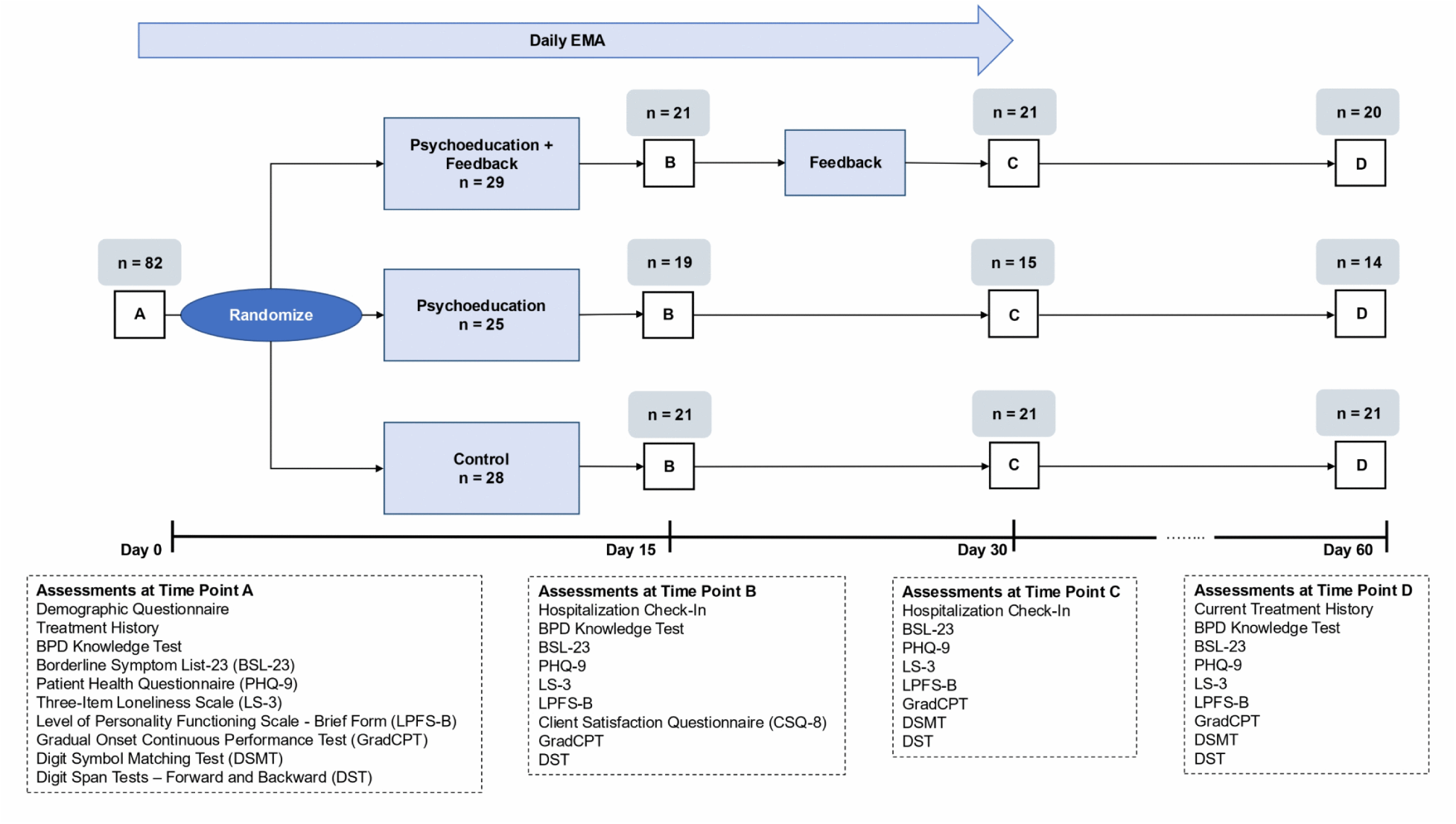
Study flow diagram.

### Measures

#### Demographics

Upon enrollment, participants reported age, ethnic background, gender, relationship or marital status, highest level of education completed, and employment status. They also reported their treatment history by answering questions adapted from the Background Information Schedule [44]. The schedule of assessments is summarized in Fig 2. All assessments were sent through links via smartphone text messaging.

#### Safety and Feasibility

We assessed the safety of our intervention using a brief hospitalization check-in questionnaire at time points B (15 days after baseline) and C (30 days after baseline). The questionnaire asked respondents whether they had experienced any of three possible events over the past 15 days: a visit to the emergency department for a psychiatric crisis, a hospitalization for a psychiatric crisis, or starting a residential psychiatric treatment program. Daily assessments included a reminder for patients that study staff would not monitor data immediately, and if they needed immediate attention or felt unsafe, they should call 911 or go to their local emergency department.

Overall feasibility was assessed in the ascertainment of the sample and retention rates in addition to a measure of acceptability for participants, the Client Satisfaction Questionnaire (CSQ-8 [45]) at time point B, the conclusion of the psychoeducational videos. In addition to asking respondents to rate their overall “satisfaction” with the service they received, the CSQ-8 solicits their impressions of the quality of service, the extent to which it matched what they were looking for, and the extent to which they would be willing to recommend the intervention to a friend or use it again themselves. The eight items are rated on a four-point Likert scale and include questions such as “Have the services you received helped you to deal more effectively with your problems?” The CSQ-8 has previously showed good internal consistency (α = .93) and validity when scores were correlated with the number of sessions attended (*r* = .56) [45].

#### Knowledge of BPD

Knowledge of BPD was measured at baseline, time point B, and time point D, using a 13-item multiple choice assessment developed by the study team based on the BPD psychoeducational video content. Questions in the first half of the survey test respondents’ knowledge of the symptoms, etiology, epidemiology, and course of BPD, while questions in the second half concern the best way for someone with BPD to manage their treatment and safety. For example, content areas assessed included the best steps to take when feeling suicidal, the extent to which medications are recommended as a component of BPD treatment, and the type of psychotherapy a person with BPD should pursue. Items were questions such as “What causes BPD?” and “What happens in the long-term course of BPD?” Cronbach’s alpha indicated suboptimal internal consistency across all timepoints (time point A: α = 0.50; time point B: α = 0.61; time point D: α = 0.62). Although reliability increased slightly over time, values remained below conventional thresholds for acceptable internal consistency. Cronbach’s alpha assumes one-dimensionality and item homogeneity. Violations of these assumptions, such as those common in knowledge-based assessments with heterogeneous content, can result in lower alpha values without necessarily indicating poor measurement quality [48]. This measure was designed to assess knowledge of commonly known facts about BPD in addition to material specific to the prescription videos. It was not designed to measure a specific construct which would lend to greater internal consistency. See S2 File for a copy of this assessment which we have name the Patient Knowledge Test-Borderline Personality Disorder (PKT-BPD; Choi-Kain, Branas, Croci, Jurist, 2022).

#### BPD Symptoms

We assessed our primary outcome of interest, overall BPD symptom severity, using the Borderline Symptom List 23 item version (BSL-23 [40]), which assesses the severity of BPD on a five-point Likert scale at baseline and time points B, C, and D. The BSL-23 is an abbreviated assessment derived from the 95 item Borderline Symptom List [46]. Sample items include “My mood rapidly cycled in terms of anger, anxiety and depression” and “Criticism had a devastating effect on me.” The measure shows high internal consistency (α = 0.935-0.969) [40]. In addition, the scale has good validity with a large effect size (*d* = 0.47), differentiating between patients with BPD and those without BPD [47]. A composite risk variable was created by summing the NSSI and suicide attempt items from the BSL-23 Supplement. Items were rated on a 5-point scale ranging from 0 (“Not at all”) to 4 (“Daily or more often”), with higher composite scores reflecting greater frequency of these behaviors.

#### Secondary Outcomes

Secondary clinical outcomes of interest were depression and personality dysfunction, assessed using the Patient Health Questionnaire (PHQ-9 [41]) and Level of Personality Functioning Scale-Brief Form (LPFS-BF [43]), respectively. The PHQ-9 is a nine-item measure of frequency of depression symptoms over the past two weeks measured on a four-point Likert scale. Sample items include “little interest or pleasure in doing things” and “feeling down, depressed, or hopeless”. The measure has strong internal reliability (α = .89) and strong construct validity, determined by comparing PHQ-9 scores with mental health professionals’ ratings of functional status and symptom-related difficulty [41]. The LPFS-BF [43] is a 12-item measure that uses a four-point Likert scale to examine the severity of personality disorder symptoms as it relates to functional impairment, yielding a two-factor solution mapping onto self and interpersonal domains [43]. Examples of items on the self-scale include, “I often think negatively about myself”, “I often do not know who I really am,” and “I often do not understand my own thoughts and feelings.” Examples of items on the interpersonal subscale include, “I often do not fully understand why my behavior has a certain effect on others” and “I often do not succeed in working cooperatively with others in an equal way.” The LPSF-BF has demonstrated acceptable reliability (α = .69), sufficient internal consistency in psychometric studies [43], convergent validity with the Severity Indices of Personality Problems Short Form (SIPP-SF [49–50]), and sensitivity to change [51]. The LPFS-BF is also more sensitive to detecting change in response to treatment than assessment of specific personality traits [52].

Additional clinical measures collected included the Three-Item Loneliness Scale [42]. This brief scale was derived from the 20-item R-UCLA Loneliness Scale [53] and contains items such as “How often do you feel left out?” on a three-point Likert scale. The scale shows fair reliability (α = .72) and good validity with the correlation with the longer R-UCLA Loneliness scale being high (*r* = .82, *p* < .001) [42].

#### Ecological momentary assessment

On each of the first 30 days of their participation in the study, participants were sent a link via text messaging to complete a short survey about their behavior and emotional experiences over the past 24 hours and a brief battery of cognitive assessments. These surveys primarily consisted of broad behavioral questions to test the feasibility of collecting information relevant to understanding respondents’ interpersonal hypersensitivity. Specifically, we asked whether they had attended work or school that day, how much time they had spent alone since completing the last survey, how many times they interacted with someone significant in their life, whether they had experienced any stressful interactions, and whether they had experienced any supportive connections. When a participant endorsed experiencing a stressful interaction or supportive connection, they were then asked a series of multiple-choice follow-up questions in which they categorized the type of interaction, characterized their emotional response to it, and reported whether they had engaged in certain high-risk behaviors since the interaction.

The daily cognitive assessments following this behavioral survey assessed response inhibition and working memory using abbreviated versions of the Gradual Onset Continuous Performance Test (GradCPT [54]) and the Digit Symbol Matching Test (DSMT [55]). These tests were administered in full in the longer batteries of assessments at the study time points.

#### Cognitive testing

Cognitive tests were administered across the four time points using the Test My Brain (TMB) platform [55–56], except for the DSMT, which was excluded at time point B. The time points at which each test was administered are depicted in Fig 2. Abbreviated versions of two of these tests, the GradCPT and the DSMT, were included in the daily EMA described above.

The GradCPT, a measure of response inhibition, sustained attention, and cognitive control, displays a series of images on a screen and asks participants to press a button when a city appears and refrain from doing so when a mountain appears. Between 80% and 90% of the images are cities, and only 10% to 20% are mountains. D′ (d-prime) is a signal detection theory metric used to quantify an individual’s ability to discriminate targets from non-targets. It is calculated as the difference between the z-transformed hit rate and the z-transformed false alarm rate with higher values indicating better discrimination accuracy [54].

In the Digit Span Test – Forward and Backward (DST), participants are shown lists of digits and asked to recall them either in the same (forward condition) or reverse (backward condition) order of their original presentation. A high score in the forward condition (a higher number of digits recalled) represents stronger attention and concentration ability; a high score in the backward condition indicates effective working memory. The DSMT is a measure of cognitive processing speed and visual short-term memory. Participants are given a key of symbols and numbers that correspond to each other for the purpose of the test and then tasked with matching symbols to numbers for 90 seconds. Scores are based on number of items completed and accuracy.

#### Statistical methods

All analyses were conducted in R (version 4.5.2) under the intention-to-treat principle [57]. Study investigators were blinded to group assignments until after the analysis of primary outcomes. All tests were two-sided with α = 0.05. Baseline characteristics were summarized using means and standard deviations for continuous variables and frequencies (%) for categorical variables. Between-group differences at baseline were evaluated using one-way analysis of variance (ANOVA) for continuous variables and Chi-square tests for categorical variables.

Safety and acceptability outcomes were examined separately at each assessment time point. Continuous outcomes were analyzed using one-way ANOVA and categorical outcomes using Fisher’s exact tests.

Primary longitudinal outcomes included BPD knowledge and overall BPD symptom severity. Secondary symptomatic measures assessed included the LPFS total score and subscores, PHQ-9, TMB-D-prime, and LS-3. Change scores at follow-up time B, C, and D relative to baseline were the main outcomes. Because longitudinal outcomes exhibited non-monotone missingness, multiple imputation with 10 imputations and 50 iterations by chained equations was used. Since symptom severity may influence study engagement, a no self-censoring missing-not-at-random mechanism [58–59] was assumed, under which missingness may depend on observed and missing symptoms. Regression analyses were conducted within each imputed dataset with treatment group modeled as a categorical predictor of change scores. Estimates and standard errors were pooled using Rubin’s rules [60]. Effect sizes for each contrast were calculated using Cohen’s *d*.

Group differences and temporal trend in EMA measures were analyzed using mixed-effects models to account for repeated observations within individuals. Ordinal outcomes (frequency of social interaction and time spent alone; four levels) were analyzed using cumulative logit mixed models with random intercepts [61]. Continuous EMA outcomes (negative affect and behavioral measures) were analyzed using linear mixed-effects models with random intercepts. All models included fixed effects for time and the group-by-time interaction. To link EMA-derived time changes in BPD symptoms to social context, we examined the association between change in BSL scores at time point C relative to baseline and time spent alone. For this analysis, we computed each participant’s mean level of time spent alone across observations, coding the ordinal responses as integers from one to four. This participant-level summary was then analyzed using linear regression, with mean time spent alone, group, and their interaction included as predictors.

The binary indicator of stressful interactions was analyzed separately. Temporal dependence was assessed by testing lagged associations using a panel model with individual-level random effects [62]. Concurrent associations among stressful interactions, negative affect, and behavior were examined using linear mixed-effects models including both random intercepts and random slopes for stressful interactions.

## Results

### Sample and Demographics

Of the 142 participants screened, 82 ultimately enrolled and completed the baseline survey. Twenty-seven participants (32.9%) did not complete the final time point and were classified as dropouts. Fig 1 provides further detail on recruitment sources and reasons for exclusion. Twenty-eight, 25, and 29 participants were assigned to the control, psychoeducation, and psychoeducation + feedback groups, respectively. Groups were matched in demographic and baseline features with a mean age of 26.8 (SD = 8.77) years and predominantly female (81.5%) and white (65.4%). Most participants had attended some college (44.4%) or completed a bachelor’s (27.2%) or graduate degree (13.6%). Chi-square tests showed no statistically significant differences between the three groups at baseline in their gender, ethnicity, or educational attainment. For further detail on the demographics of the sample, see Table 1. At baseline, one-way ANOVAs showed no significant baseline differences in BPD symptoms, depression, personality dysfunction, or knowledge of BPD.

**Table 1.** Sample characteristics by treatment allocation.

|  |  | Total | Control | Psychoed | Psychoed + Feedback |
| --- | --- | --- | --- | --- | --- |
| Number of participants |  | 82 | 28 | 25 | 29 |
| Mean age, years (SD) |  | 26.8 (8.77) | 27.7 (11.7) | 26.4 (6.97) | 26.3 (6.87) |
| Gender (female) |  | 67 (81.7%) | 25 (89.3%) | 19 (76.0%) | 23 (79.3%) |
| Ethnicity | African/Black | 5 (6.1%) | 3 (10.7%) | 1 (4.0%) | 1 (3.4%) |
|  | American Indian or Alaska Native | 1 (1.2%) | 0 (0%) | 1 (4.0%) | 0 (0%) |
|  | Asian | 8 (9.8%) | 4 (14.3%) | 3 (12.0%) | 1 (3.4%) |
|  | White | 53 (64.6%) | 17 (60.7%) | 16 (64.0%) | 20 (69.0%) |
|  | Other | 6 (7.3%) | 4 (14.3%) | 1 (4.0%) | 1 (3.4%) |
|  | Multiple | 9 (11.0%) | 0 (0%) | 3 (12.0%) | 6 (20.7%) |
| Education | HS or lower | 12 (14.6%) | 5 (17.9%) | 2 (8.0%) | 5 (17.2%) |
|  | Some college | 37 (45.1%) | 11 (39.3%) | 15 (60.0%) | 11 (37.9%) |
|  | Bachelor's degree | 22 (26.8%) | 8 (28.6%) | 5 (20.0%) | 9 (31.0%) |
|  | Graduate degree | 11 (13.4%) | 4 (14.3%) | 3 (12.0%) | 4 (13.8%) |
| **Baseline Symptoms** |  |  |  |  |  |
| BSL-23 | Mean (SD) | 2.28 (0.847) | 2.23 (0.884) | 2.17 (0.855) | 2.43 (0.813) |
|  | Median [Min, Max] | 2.39 [0.739, 3.74] | 2.17 [0.739, 3.65] | 2.22 [0.913, 3.65] | 2.48 [0.826, 3.74] |
| PHQ-9 | Mean (SD) | 16.1 (5.44) | 15.4 (6.21) | 16.0 (5.01) | 16.8 (5.10) |
|  | Median [Min, Max] | 16.0 [1.00, 26.0] | 14.5 [5.00, 26.0] | 18.0 [1.00, 26.0] | 17.0 [6.00, 25.0] |
| LPFS-BF | Mean (SD) | 35.4 (6.01) | 34.1 (7.37) | 36.6 (5.17) | 35.7 (5.11) |
|  | Median [Min, Max] | 35.5 [21.0, 48.0] | 35.0 [21.0, 47.0] | 36.0 [24.0, 48.0] | 35.0 [23.0, 47.0] |
| BPD Knowledge | Mean (SD) | 9.94 (1.98) | 9.89 (2.06) | 9.28 (2.25) | 10.6 (1.48) |
|  | Median [Min, Max] | 10.0 [4.00, 13.0] | 10.0 [4.00, 13.0] | 9.00 [6.00, 13.0] | 11.0 [7.00, 13.0] |
| BSL-23 Supplement 1 | Mean (SD) | 0.443 (0.663) | 0.571 (0.836) | 0.387 (0.567) | 0.368 (0.544) |
|  | Median [Min, Max] | 0.167 [0, 4.00] | 0.333 [0, 4.00] | 0 [0, 2.00] | 0 [0, 2.33] |
| BSL-23 Supplement 2 | Mean (SD) | 0.477 (0.443) | 0.469 (0.526) | 0.495 (0.345) | 0.470 (0.445) |
|  | Median [Min, Max] | 0.375 [0, 2.63] | 0.375 [0, 2.63] | 0.500 [0, 1.13] | 0.375 [0, 1.50] |

BSL-23 = Borderline Symptom List – Short Version, PHQ-9 = Patient Health Questionnaire-9,
LPFS-BF = Level of Personality Functioning Scale-Brief Form, BSL-23 Supplement 1 =
Borderline Symptom List Supplement: Items for Assessing Behavior Self-Harming Behaviors,
BSL-23 Supplement 2 = Borderline Symptom List Supplement: Items for Assessing Behavior
High-Risk Behaviors.

The majority (56.77%) of participants reported no prior exposure to specialized treatment, and the sample, on average, had 2.02 (*SD* = 2.57) years of either individual or group therapy. There were no significant between group differences between the amount of DBT or TFP exposure, but the psychoeducation group without feedback received significantly more MBT (*N* = 3) than the other two groups, which received none. Participants indicated, on average, that they had first received psychiatric treatment 7.68 years ago (*SD* = 7.52), despite only recently receiving their BPD diagnosis.

### Safety and Acceptability

Safety was assessed based on self-reported visits to the emergency room, hospitalization, and step up to residential care. The frequencies of these events were compared across the three groups using Fisher’s exact tests. Results indicated no significant group differences in any of the three safety measures at either time point B or time point C (Table 2). Overall, these events were infrequent: within the first 30 days of the study, there were two emergency department visits, eight hospitalizations, and one transition to residential level of care.

**Table 2.** Emergency department visits, hospitalizations, and entrance of residential treatment at two time points following psychoeducational intervention.

|  |  | Total |  | Control |  | Psychoed |  | Psychoed + Feedback |  | *p* Value |
| --- | --- | --- | --- | --- | --- | --- | --- | --- | --- | --- |
|  |  | n | Events | n | Events | n | Events | n | Events |  |
| ER Visit | Time B | 57 | 0 (0%) | 18 | 0 (0%) | 18 | 0 (0%) | 21 | 0 (0%) | NA |
|  | Time C | 53 | 2 (3.77%) | 21 | 0 (0%) | 13 | 1 (8.33%) | 19 | 1 (5.56%) | 0.512 |
| Hospitalization | Time B | 57 | 8 (14%) | 18 | 3 (16.7%) | 18 | 3 (16.7%) | 21 | 2 (9.52%) | 0.714 |
|  | Time C | 53 | 0 (0%) | 21 | 0 (0%) | 13 | 0 (0%) | 19 | 0 (0%) | NA |
| Residential | Time B | 57 | 0 (0%) | 18 | 0 (0%) | 18 | 0 (0%) | 21 | 0 (0%) | NA |
|  | Time C | 53 | 1 (1.89%) | 21 | 0 (0%) | 13 | 0 (0%) | 19 | 1 (5.26%) | 0.604 |

Dropout rates were used as an indicator of intervention acceptability. Over the course of the study, 32.9% of participants failed to complete a scheduled assessment and did not complete any subsequent time points; these individuals were classified as dropouts. Dropout rates did not differ significantly across groups (control = 25%; psychoeducation = 44%; psychoeducation + feedback = 31%; *p* = .34), and dropout rates were similar across time points (*p* = .12).

Client satisfaction was assessed using the CSQ-8 [45]. Participants in the psychoeducation groups who received BPD-related videos reported a mean score of 24.0 (*SD* = 4.64), compared to a mean score of 20.6 (*SD* = 5.36) in the control group, after viewing their assigned videos. The one-way ANOVA indicated that satisfaction was significantly higher in the BPD psychoeducation groups than in the control group (*p* = .016).

### Change in BPD Knowledge

Analyses indicated no floor effects at any timepoint using this newly developed measure of BPD knowledge, suggesting that the measure was not too difficult for participants and that minimum scores were not reached. In contrast, ceiling effects were observed and increased over time, indicating that a growing proportion of participants achieved the maximum possible score at follow-up assessments. See Table S2 for the change for each item across condition and time.. When examined by condition, the control group showed a modest ceiling effect at baseline (10.7%) that was not observed at later timepoints, suggesting stable and relatively limited BPD knowledge. The psychoeducation group demonstrated minimal ceiling effects at baseline (4%), which increased at time point B (27.8%) before decreasing at time point D (14.3%), indicating an initial increase in knowledge followed by a reduction in scores over time. The psychoeducation with feedback group showed the strongest ceiling effects, increasing from 3.45% at baseline to 28.6% at time point B and 35% at time point D, suggesting substantial knowledge gains throughout the study.

Average levels of BPD knowledge increased over the course of the study at time points B and C for both psychoeducation groups, while BPD knowledge decreased in the control condition after the generic mental health video intervention. Only the psychoeducation with feedback condition showed a steady increase in average levels of BPD knowledge at each subsequent time point (Table 3). The pooled regression results across multiple imputations showed that both groups receiving BPD-related education, psychoeducation (β = 1.65, SE = 0.72, *p* = .03) and psychoeducation + feedback conditions (β = 1.16, SE = 0.67, *p* = .09), displayed a larger increase in knowledge after receiving the video psychoeducation for two weeks compared to the control group. Of note, the psychoeducation group had more patients exposed to some MBT while enrolled in the study but showed limited increase in BPD knowledge at time point C, two weeks after the video psychoeducation ended.

**Table 3.**
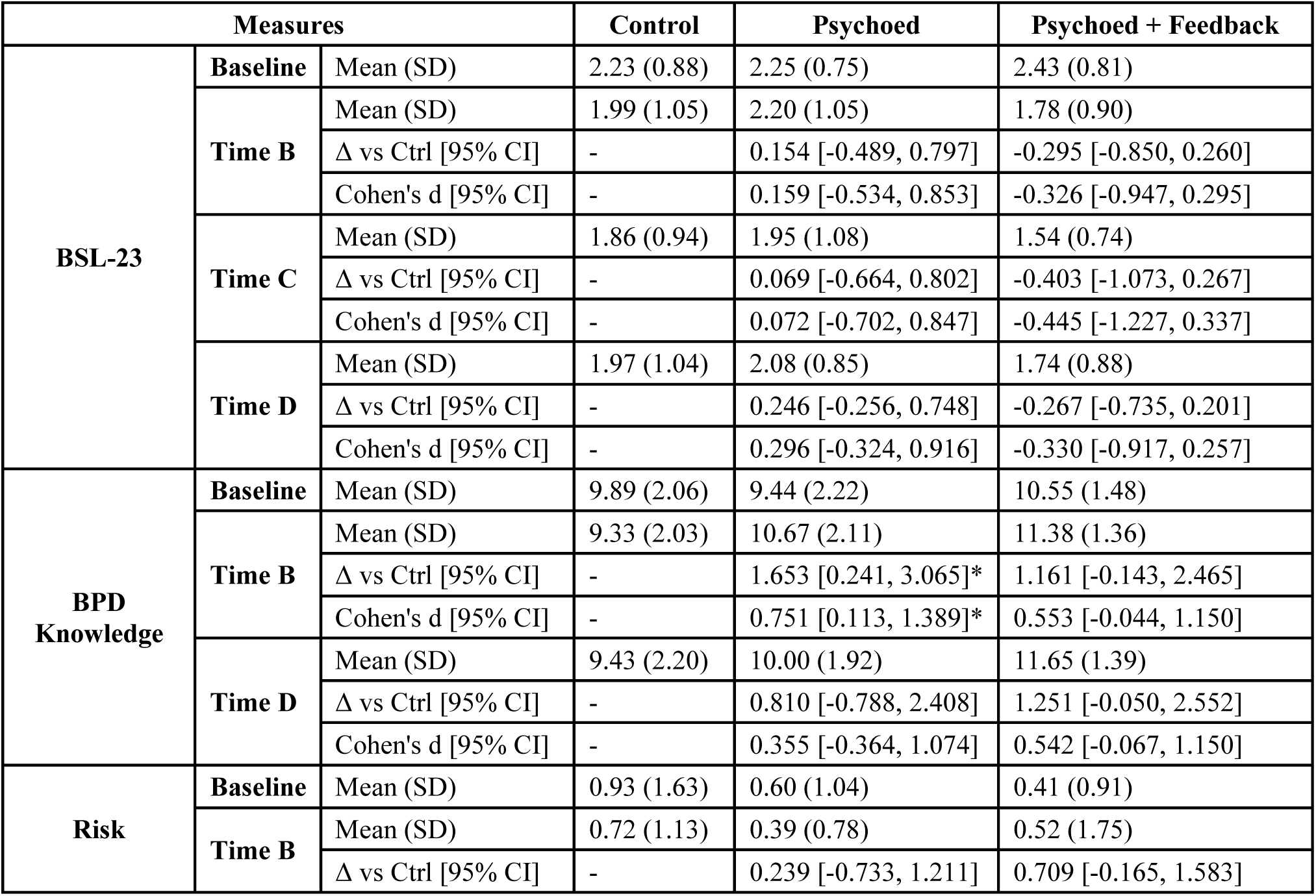
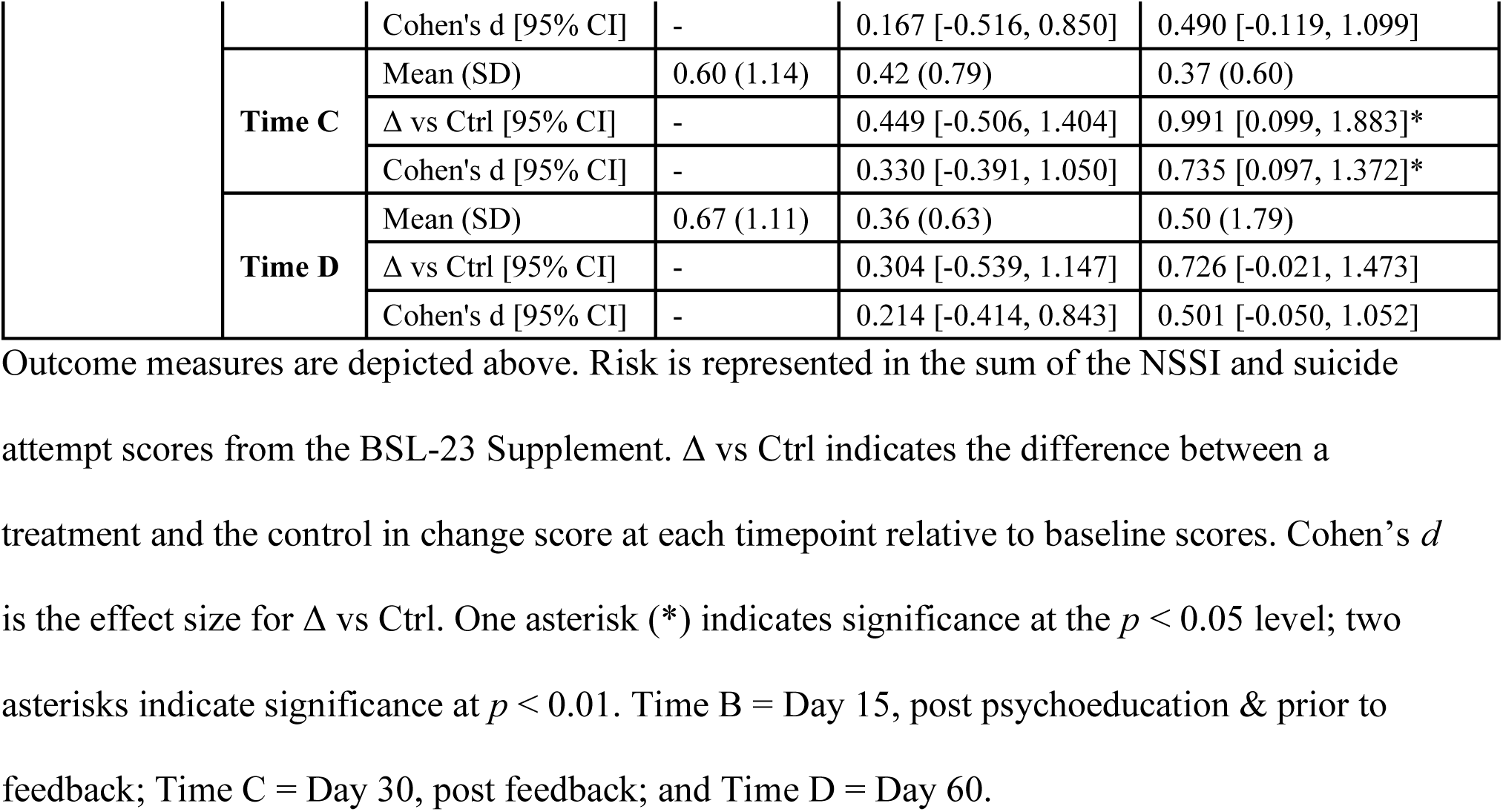
Primary outcome measures by group at baseline and at the three follow-up time points.

### Change in Overall BPD Symptoms

BSL scores declined for the control and psychoeducation + feedback group at time points B, C, and D relative to baseline (Fig 3). In the psychoeducation group, there was close to zero change at time point B, and a slight increase in BSL scores at time point D. The psychoeducation + feedback showed the largest improvement in overall BPD symptoms across the three time points, but the difference in that change from baseline was not significant (*p* = .27) compared to the other two groups (Table 3). Descriptive statistics for the composite risk variable derived from the BSL-23 Supplement are presented below (Table 4). A linear mixed effects model with random intercepts for participant revealed that neither time (*F* = 0.42, *p* = .641), condition (*F* = 0.32, *p* = .878), nor the time by condition interaction (*F* = 1.14, *p* = .34) significantly predicted risk. In addition, a Poisson mixed effects model demonstrated that baseline risk did not significantly predict dropout (β = -0.011, *p* = .546).

**Fig 3.**
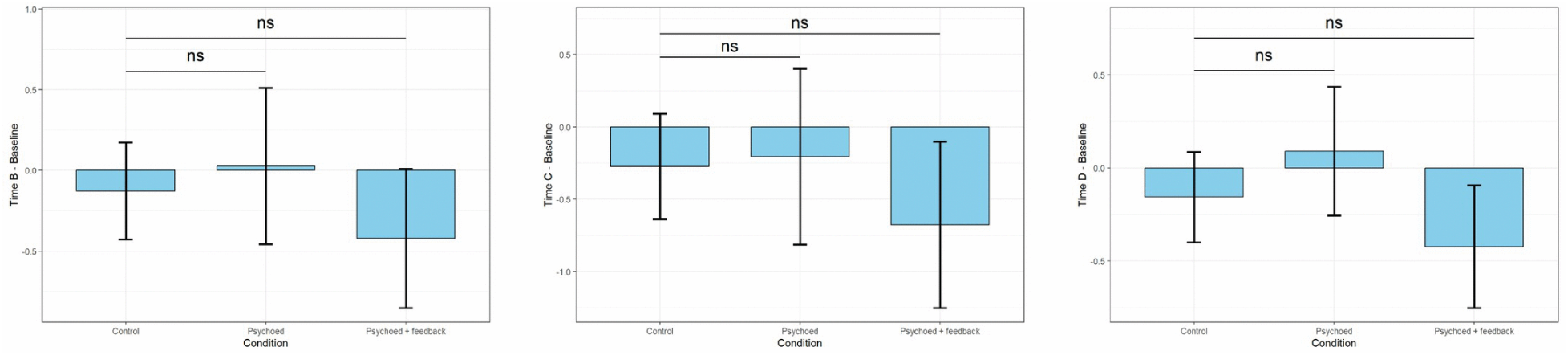
Estimated mean change in BSL-23 scores at time points B, C, and D by treatment group (control, psychoeducation, psychoeducation + feedback). Blue bars represent model-estimated means and error bars indicate 95% confidence intervals. Pairwise group comparisons are shown above each panel; ns = not statistically significant at significance level 0.05.

**Table 4.** Descriptive Statistics for the Composite Risk Variable.

|  |  | Total | Control | Psychoed | Psychoed + Feedback |
| --- | --- | --- | --- | --- | --- |
|  |  | *n* | *Mean (SD)* | *Mean (SD)* | *Mean (SD)* |
| **NSSI & Suicide Attempt** | **Baseline** | 82 | 0.93 (1.63) | 0.60 (1.04) | 0.41 (0.91) |
|  | **Time B** | 57 | 0.72 (1.13) | 0.39 (0.78) | 0.52 (1.75) |
|  | **Time C** | 51 | 0.60 (1.14) | 0.42 (0.80) | 0.37 (0.60) |
|  | **Time D** | 55 | 0.67 (1.11) | 0.36 (0.63) | 0.50 (1.79) |

Items were rated on a 5-point scale: 0 (“Not at all”), 1 (“Once”), 2 (“2–3 times”), 3 (“4–6 times”), and 4 (“Daily or more often”), with higher composite scores reflecting greater frequency of these behaviors.

### Change in Secondary Symptomatic Measures

Results for the secondary symptomatic outcomes indicated no significant group differences in change from baseline in PHQ-9, LPFS total score, or LPFS interpersonal score at time points B, C, or D (Table 5, all *p* > .10). However, participants in the psychoeducation + feedback group showed a significantly greater reduction in LPFS self-score at the final time point compared with the control group (β = −1.82, *p* = .02) with a significant medium effect size of 0.659, indicating greater improvement in self-functioning in the psychoeducation + feedback condition at this time point. Changes of loneliness (LS-3) relative to baseline did not differ significantly across groups at any follow-up time points. D-prime, a measure of attentional control and capacity to differentially response to target versus non-target stimuli, on average increased steadily over the timepoints in the psychoeducation + feedback group but showed no consistent trend in the other groups. However, these changes in cognitive control showed no significant group differences (Table 5, all *p* > .40).

**Table 5.**
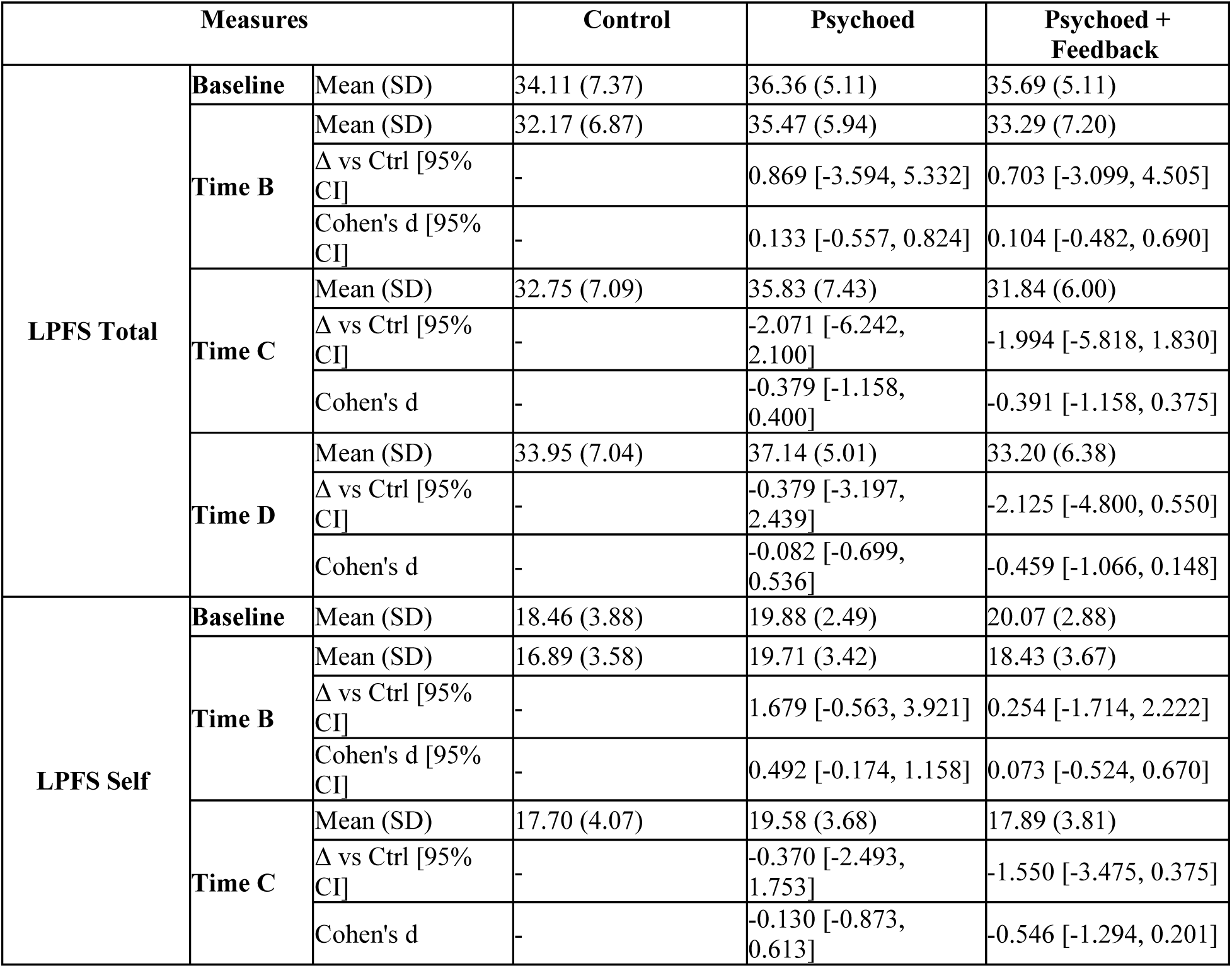
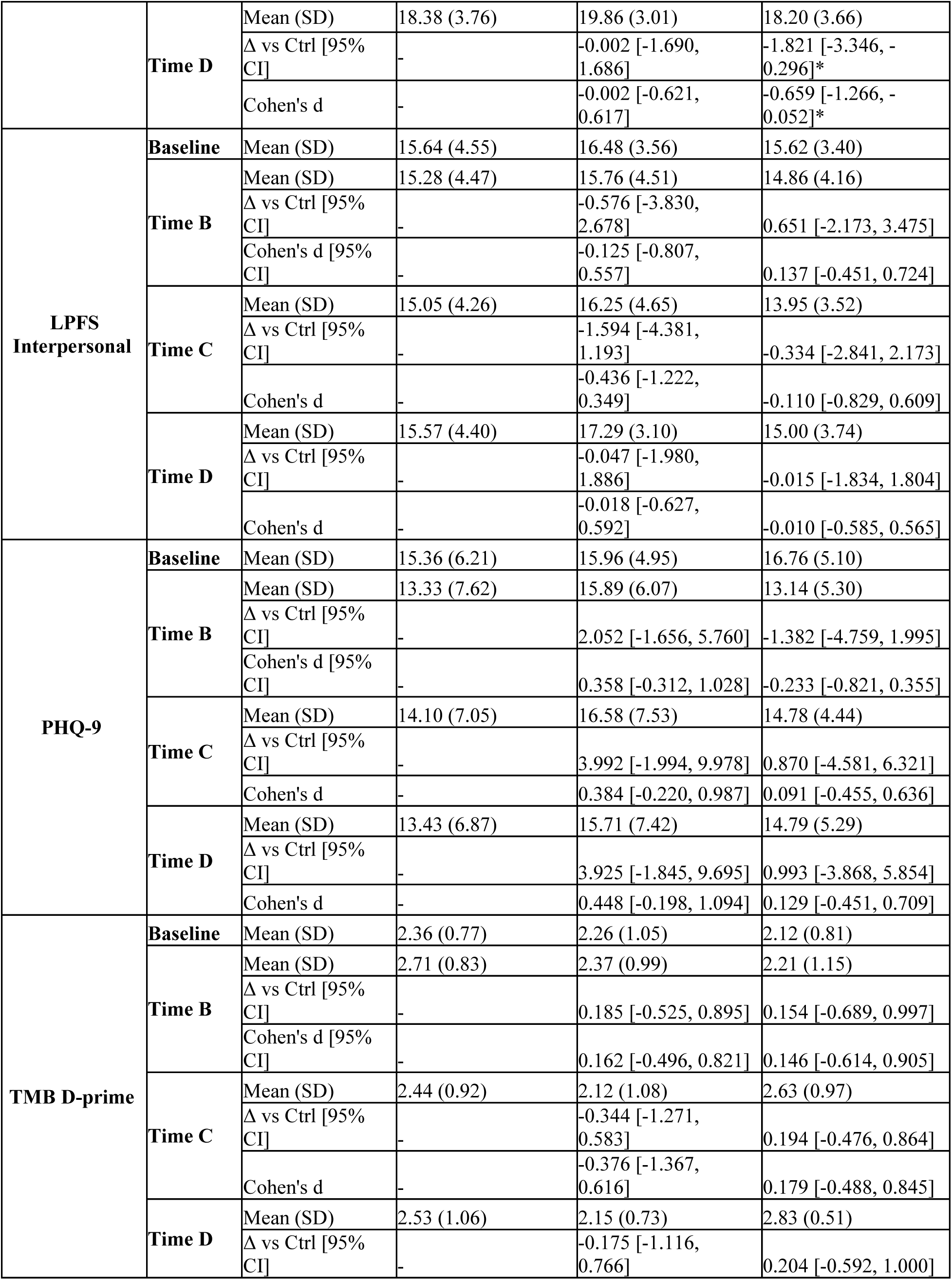
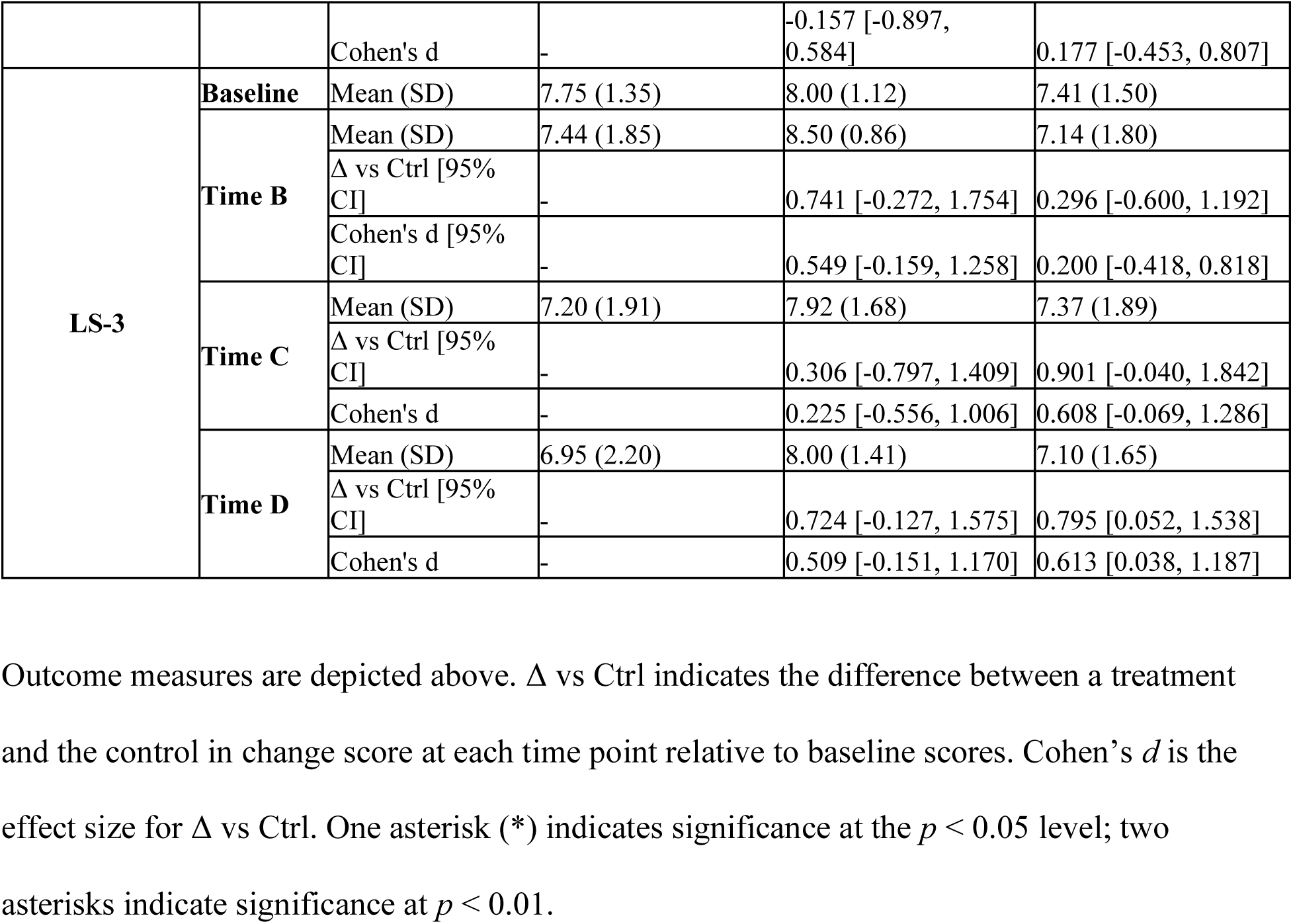
Secondary Outcome Measures for Each Group at time points B, C, and D.

### Results from EMA

The temporal trends and their differences across groups of several EMA measures were estimated with mixed-effect models. For time spent alone, the model showed no significant overall effect of time (β = -0.01, *p* = .213) and no significant baseline differences between either the psychoeducation group (β = -0.30, *p* = .579) or the psychoeducation + feedback group (β = - 0.76, *p* = .131) compared with the control group. However, significant time-by-group interactions were observed for both the psychoeducation group (β = 0.04, *p* = .013) and the psychoeducation + feedback group (β = 0.07, *p* < .001), indicating that the odds of having more time spent alone increased over time in both intervention groups, with a stronger effect in the psychoeducation + feedback group. For number of interactions, there was a significant overall decrease over time (β = -0.02, *p* = .003) in the control group. The psychoeducation group did not differ from control at baseline (β = -0.09, *p* = .829), whereas the psychoeducation + feedback group reported more interactions than control at baseline (β = 1.03, *p* = .014). Over time, no differential temporal trend was observed for the psychoeducation group (β = -0.00, *p* = .734). However, the psychoeducation + feedback group showed a greater decline in number of interactions over time (β = -0.02, *p* = .036). Negative affect decreased slightly over time (β = - 0.10, *p* = .048) in the control group. There were no significant baseline differences in negative affect between groups and no significant time-by-group interactions, indicating that changes in negative affect over time did not differ by treatment group. Similar to negative affect, there was no overall effect of time on impulsive behavior (β = 0.00, *p* = .973) and no significant time-by-group interactions. The psychoeducation group showed slightly more impulsive behavior at baseline compared to control (β = 0.40, *p* = .048), whereas the psychoeducation + feedback group did not differ from control.

In the model examining the association between changes in BSL-23 scores at time point C and mean time spent alone, no significant main effects of condition were observed. In the control group, mean time alone showed a marginal positive association with change in BSL-23 (β = 0.43, *p* = .077), indicating more time spent alone was correlated with more severe BPD symptoms. On the other hand, the interaction between mean time alone and the psychoeducation + feedback group was significant and negative (β = −0.66, *p* = .047), which means for patients receiving psychoeducation + feedback, more time spent alone was correlated with improved BPD symptoms. The psychoeducation group exhibited a similar association structure as the control group since the interaction term for this group was not significant.

Autoregressive analysis of stressful interactions resulted in a positive coefficient (*r* = 0.252, *p* < 0.001), demonstrating that having stressful interactions one day increased the likelihood of having stressful interactions the next day. Mixed-effects models indicated significant positive concurrent associations between stressful interactions and both negative affect and impulsive behavior (β = 0.24, *p* < .001), suggesting that the occurrence of a stressful interaction was associated with elevated negative affect and greater impulsive behavior within the same day.

## Discussion

This pilot study evaluating the safety and feasibility of online psychoeducation and assessment as a first step of care for newly diagnosed BPD yielded four main findings: 1) online psychoeducation and assessment integrating EMA with feedback is safe, feasible, and acceptable to participants with newly diagnosed BPD; 2) online psychoeducation about BPD resulted in both higher client satisfaction and BPD knowledge levels than the control group; 3) the psychoeducation with feedback condition was associated with consistent decreases in BPD symptoms at each time point but was not superior to the other conditions at a statistically significant level; and 4) the psychoeducation with feedback condition showed greater improvements in self-impairment in personality functioning than the other conditions at the final time point.

Two additional findings regarding the relationship between daily social interactions and BPD symptoms provide testing of the interpersonal hypersensitivity model. First, using the EMA data collected across the entire study, we found stressful interpersonal interactions were associated with both concurrent negative affect and impulsivity, as well as increased likelihood of having another stressful interaction the next day, highlighting the link between symptomatic domains and social stress. Perhaps our most notable finding is that in the control group, which showed a pattern of decline in BPD knowledge over time and minimal change in self-impairment scores, we uncovered an association between increases in BPD symptoms and more time spent alone. In contrast, the psychoeducation with feedback group, which demonstrated increases in BPD knowledge over the four time points and significantly greater reductions in self-impairment scores than the control group at the end of the study, showed an inverse association between BPD symptoms and time spent alone.

Altogether, these results confirm the safety and feasibility of a brief virtual intervention to provide a first step of care with psychoeducation, assessment, and feedback following diagnostic identification of BPD in many clinical contexts where availability of other care that meets national guidelines is typically not immediately available. Preliminary signs of the effectiveness of psychoeducation combined with feedback--increasing BPD knowledge while decreasing self-impairment and BPD symptoms when alone—were found in contrast to general psychoeducation where these outcomes were absent.

Regarding feasibility, we ascertained an adequate sample for randomization but encountered several obstacles in recruitment and did not reach our goal of 100 participants [39]. Most participants who dropped out (20 of 27) were those who had been recruited from inpatient units, who were likely more symptomatically acute than those recruited online. In general, those who dropped out did so early in the study, with eight of the 27 dropouts occurring immediately after baseline (i.e., no daily surveys or subsequent time points were recorded) and all but five occurring before time point B. Our dropout rate of 32.9% is consistent with similar online BPD intervention studies, which ranges from 0 to 56.7% (*M* = 22.5 [63]), as well as the rates of dropout in RCTs of 28.2% derived from a meta-analysis of BPD-related intervention studies [64]. Regarding safety, no adverse events were reported to study staff. Eleven (19.2%) participants reported entering a higher level of care during the study with no differences between treatment arms. This rate of stepping up in level of care approximates previously reported readmission rates to inpatient hospitals for individuals with personality disorders [65]. Additionally, we calculated risk using the items participants self-reported regarding self-harm and suicide attempts from the BSL-23 supplement, finding no significant increase in risk over time as well as no difference is self-destructive behavior between the three arms of the study.

Our second finding is that those who received psychoeducation about BPD, in contrast to those receiving matched high quality general mental health psychoeducation, reported both higher client satisfaction and significantly greater improvements in knowledge about BPD. These findings illustrate both acceptability to users and effectiveness of BPD psychoeducation using brief online video prescriptions educating patients about their condition.

While our pilot study was not adequately powered to assess effectiveness, preliminarily, we found that BPD symptoms declined for all groups over the course of the study. In the psychoeducation group, there was close to zero change after videos and a slight increase in symptoms by the end of the study, despite a significant increase in BPD knowledge after viewing videos. We found that the magnitude of improvements in BPD symptoms appears greatest in the psychoeducation with feedback arm, which decreased from a mean of 2.43 at baseline to 1.78 after videos and 1.54 after feedback. Only the psychoeducation with feedback group showed adequate changes in mean scores qualifying for change in severity categories from high (1.87-2.67) to moderate (1.07 to 1.87) [47]. However, we did not find statistically significant differences between groups, likely due to the underpowered sample size. Also, notable is that psychoeducation about depression, anxiety, and other mental health concerns in the control condition was not associated with greater change in depression ratings despite providing more depression-related content.

These findings may suggest that psychoeducation alone, available on widely accessible platforms such as YouTube, may not be sufficient to prescribe to patients to improve outcomes for individuals with BPD. Our BPD psychoeducation intervention involved a relatively low dose of treatment, at approximately 66 minutes in total length, compared to a matched psychoeducational intervention of similar quality, expertise, and institutional prestige about depression, anxiety, and general mental health concerns. Prior trials of psychoeducation about BPD use waitlist comparisons [19–20], unmatched control conditions [21], or digital or video intervention in addition to CAU [26, 28, 34]). To our knowledge, this is one of only two RCTs evaluating a video intervention with feedback [34] and the only one using online videos via YouTube links. Our results are consistent with the results of three of the four published RCTs on digital interventions for BPD using matched control conditions, the majority of which also reported no significant between-group differences in change in BPD symptoms [26–27]. The latest *Priovi* study, which included 580 participants with a considerably higher dose of intervention, is to date the only RCT demonstrating effectiveness of an online add-on treatment in reducing BPD symptoms with a small effect size [28]. The previously published research on *Priovi* using a smaller but still large sample of 204 participants notably did not find statistically significant differences in favor of the addition of this online intervention [26]. These studies suggest that the sample size required to detect significant differences for digital interventions is very large considering that relative to other psychopathology, BPD has a low rate of responsiveness to intensive comprehensive therapies, such as DBT [66]. BPD is a complex disorder that requires longer courses of treatment for a more definitive treatment response.

However, comprehensive intensive therapies for BPD only achieve small to moderate effect sizes in a meta-analytic review [6] with total contact hours exceeding 20 hours up to 100 hours in 12 months of treatment if considering twice weekly formats involving either group and individual therapy or individual therapy twice weekly. Interventions such as this one are not intended to replace such treatments or be comparable in their effects. While this type of brief first step of care will likely have small effects, psychoeducation and assessment will have a clinically important impact, which on a broader population and systems level may have effects that a pilot RCT cannot detect. Furthermore, components that inform patients about their illness and that meet guidelines-based standards should be embraced if proven safe and feasible, even if effects are not obvious. Research is still needed to understand what package of feasible guidelines-based interventions can be made more accessible, while also yielding measurable clinical and empirical effects. Future studies will require a sample size that is much greater in magnitude with an increased dose of psychoeducation, assessment, and feedback.

Our fourth finding is that the psychoeducation with feedback arm was associated with significantly greater improvement in self-impairment, based on LPFS scores, by the end of the study than the control condition. The self-impairment scale on the LPFS-BF includes items related to problems of self-clarity and self-control, in addition to high negativity and unrealistic thinking regarding oneself. These items map onto the identity domain of personality disorders, which relates to experiences of “painful incoherence” of thoughts, feelings, and behavior; difficulty in commitment to jobs or roles; and role absorption, or tendency to define oneself by a single role [67]. Meares et al. [68] proposed that BPD’s central disturbance operates in the self-system where significant dependency on others emerges from an otherwise impoverished sense of self. He described self-fragmentation and failure in synthesizing a coherent understanding of one’s own experience as a factor contributing to the intolerance of aloneness [69], as well as a deficiency in higher order cognitive processes that enable mentalization, that is, the capacity to realistically and flexibility understand the mental states of oneself and others in social interactions.

Gunderson’s GPM formulation of BPD as a disturbance of interpersonal hypersensitivity identifies the intolerance of aloneness and underdeveloped self-reliance as a driver of dependency on caregivers [22–23]. This overdependency impairs the capacity to use healthcare optimally. Overdependency, as a personality trait, is associated with greater emergency room visits and somatic concerns, as well as poorer stress management, eating habits, and overall health behaviors [70]. Our detection of significant change with a medium effect size emphasizes that self-impairment is a viable and responsive target of treatment for patients with high severity personality dysfunction that may respond to routine and reliable psychoeducation with feedback. More research is needed to track treatment response in terms of personality functioning along with important measurable outcomes, such as therapeutic alliance, increasing relationship stability outside of treatment, and reduced reliance on acute healthcare services.

Our finding suggests that learning about BPD via psychoeducation with personalized feedback based on online assessment—compared to learning about general mental health concerns or psychoeducation about BPD alone— may organize and enhance self-knowledge, improving functioning in the self-system. This preliminary finding may indicate that the effect of psychoeducation may be more clinically detectable when combined with personalized feedback for improving self-clarity and painful incoherence. This finding provides a data-driven verification of what people with lived experience express about psychoeducation via YouTube videos in qualitative studies [33]. Whether the change detected in the final time point of our study reflects state-related change in level of distress or more stable change in personality functioning is not known and would require a longer period of follow-up to assess. Contrary to expectations, we did not find significant changes or differences between groups on the relationship subscale of the LPFS at any time point. Further research is needed to examine whether online psychoeducation with feedback more specifically impacts interpersonal functioning in a larger, more definitive trial following participants for a longer period where this mechanism of change can be further tested.

Lastly, beyond the assessment of safety, feasibility, acceptability, and preliminary signs of efficacy, which were the primary aims of this pilot study design, the EMA protocol employed was tested to assess the feasibility of measuring daily interpersonal processes and the types, quality, and number of social interactions encountered by participants. As expected, stressful interpersonal interactions were associated with both concurrent negative affect and impulsivity, as well as increased likelihood of having another stressful interaction the next day, highlighting the link between BPD symptom domains and social stress. A more novel finding was that the psychoeducation plus feedback group showed a significant and inverse relationship between increasing time spent alone and BPD scores, which contrasts with a direct relationship between increasing time spent alone and BPD scores in the control group. Groups did not differ in reported loneliness. Considering our finding that the psychoeducation about BPD plus feedback resulted in significantly greater improvement in self-impairment, this last finding also lends evidence to the possibility that brief, online psychoeducation about BPD with personalized feedback may improve a patient’s capacity to understand themselves more clearly and spend time alone without feeling lonelier, perhaps as an indicator of self-sufficiency and tolerance of aloneness. This finding is important considering the intervention’s coverage of the concept that BPD is a problem of interpersonal hypersensitivity, where insecurity about one’s self translates into a tendency to be reactive to the availability of others in a way that leads to threatened states when real or perceived abandonment occurs, which then segues into more impulsively self-destructive states when the person with BPD feels abandoned and alone. Control participants did not receive such psychoeducation and notably showed a pattern of more BPD symptoms as they spent more time alone.

Increasing self-clarity, BPD knowledge, and the capacity to be alone may be important in the first step of treatment after a BPD diagnosis. Future research is needed to evaluate whether these outcomes relate to clinical improvements, such as reductions in readmissions to higher levels of care, self- or interpersonally damaging behavior, or problems in engagement with outpatient care, as well as functional improvements in work, school, and other community engagements, which are vital to structuring and reinforcing healthy personality functioning.

## Limitations

There are several important limitations of our methodology and the generalizability of our findings. First, this study was conducted entirely online without systematic verification of the BPD diagnosis by a clinical professional or by a reliable semi-structured interview assessment of BPD. Participants’ average self-reported symptoms were in the high severity range at baseline suggesting likelihood of meeting diagnostic criteria if asked the same question in a semi-structured interview. Further research should implement a more rigorous means of verifying the accuracy and recency of the BPD diagnosis, as well as the more complete clinical profile of participants. A second limitation was an inability to verify how many videos each participant viewed, which would provide data on how much the video intervention influenced the outcomes reported. Third, we did not control for the amount or type of care participants were receiving concurrent with this intervention. We recruited participants who had not received full-scale DBT, MBT, or TFP but could not prohibit the initiation of treatment during their involvement in our study. Our fourth limitation is that we did not assess medications or biological treatments concurrent with the trial, which may have affected outcomes. An independent means of tracking other treatments participants engaged in concurrent with this intervention would improve future studies. Lastly, this study is limited by the no-self-censoring assumption underlying the missing data model, which cannot be empirically verified against the true missing mechanism. Further research on this intervention protocol should address these limitations to produce more reliable and generalizable findings.

## Conclusion

This study preliminarily demonstrates that online psychoeducation and assessment for BPD is feasible, safe, and possibly effective in reducing self-impairment and the capacity to spend more time alone without an escalation of symptoms but only when personalized feedback is provided to improve the self-awareness individuals have about the status of their unique clinical trajectory. Free use of publicly available videos online alone might not result in detectable clinical change, even when those videos are produced by academic centers of expertise. More research is needed to further assess the effects of individual assessment and feedback of a personal clinical profile over time for added personalization of a BPD focused resource. Our results suggest that the combination of online psychoeducation with feedback is acceptable to patients and can be implemented with limited training or clinical facetime by professionals. It is expectable based on prior research on digital interventions for BPD that symptom change in the context of a study design with a matched control condition is likely to be small. No other study to our knowledge, to date, has tested a psychoeducation intervention for BPD against a matched control condition with high quality content to enhance mental health literacy. These findings advance our understanding that the use of routine systematic data collection at regular intervals with personalized measurement-based feedback, in combination with brief but high-quality video prescriptions of psychoeducation, may provide modest, isolated changes that may be clarifying and empowering for people immediately after they receive their BPD diagnosis. Providing psychoeducational video prescriptions immediately, in a way that is scalable, brief, and requires little training for frontline clinicians making the diagnosis. Considering the broadly common status quo that psychotherapy is often not the first step of care, even in the most robust of mental healthcare systems, these prescriptions offer a safe, and feasible entry to care that may be organizing for the patient in their treatment decision-making.

Assessment of key mechanisms, such as interpersonal hypersensitivity, overdependence on others, self-confusion, and incapacity to be alone, may provide meaningful insight into how change occurs in response to treatment and how they relate to both symptomatic and functional outcomes. It makes sense that educating patients with BPD about the nature of their illness, basic ways of managing safety, and recommended treatments while also providing simple feedback about individual symptom profiles, increases clarity about oneself. Self-clarity can empower those with the identity confusion that characterizes BPD to form clear goals, think first before acting, and manage stress alone. Whether or not self-clarity and the capacity to be alone translates into being less dependent on treatment, as measured by utilization of care, and more engaged in life and identity building endeavors, such as school, work, and home management, should be assessed in outcomes studies. Future studies may consider a greater dose and duration of psychoeducation with a longer period of assessment and continuous feedback, combined with a more controlled TAU, to assess whether this particular step of care enhances both symptomatic and functional outcomes, as well as their mechanisms of change. People with BPD are shown to respond to a variety of different treatments [6]. Public health interests call for prioritization of developing practical and feasible basic interventions that can be more widely implemented to reduce the morbidity and mortality, as well as the personal and societal costs of this condition, as a first step of newly diagnosed BPD.

## Data Availability

Data cannot be shared publicly because it was not allowed by the Mass General Brigham IRB. However, data is available upon request, and the MGB IRB can be contacted at for data sharing requests or questions. The IRB identification number for this study is 2022P000892.

## Acknowledgements

We would like to acknowledge Ethan Kahn for aiding in data collection and Kaitlyn Tsai for research support in the early drafting stages of this manuscript. In addition, we thank Rebbie Ratner and *BorderlinerNotes* team for producing the BPD psychoeducation videos, and our anonymous lived experience experts for feedback on our video prescriptions.

## Supporting information

**S1 Table. Psychoeducation Video Content.**

**S2 Table. Percentage Correct and Change for BPD Knowledge Items by Condition and Time Point.** 𝛥% represents absolute percentage-point change, defined as the percentage correct at time point B minus the percentage correct at baseline (or time point B for 𝛥%D).

**S3 Table. Comparison of people exposed to some DBT, MBT, or TFP versus people not exposed to any DBT, MBT, or TFP in terms of primary outcomes.**

**S1 File. Example of Feedback.**

**S2 File. Patient Knowledge Test – Borderline Personality Disorder (PKT-BPD).**

